# Two-year clinical outcomes of staged transcatheter mitral edge-to-edge repair after transcatheter aortic valve replacement

**DOI:** 10.1101/2024.03.28.24305039

**Authors:** Takashi Nagasaka, Vivek Patel, Ofir Koren, Alon Shechter, Tarun Chakravarty, Wen Cheng, Hideki Ishii, Hasan Jilaihawi, Mamoo Nakamura, Raj R. Makkar

## Abstract

**Background:** Residual significant mitral regurgitation (MR) can increase the risk of adverse events following transcatheter aortic valve replacement (TAVR). The clinical benefits of staged transcatheter edge-to-edge repair (TEER) post-TAVR remain underexplored. This study aimed to investigate clinical outcomes of staged TEER for residual significant MR post-TAVR.

**Methods:** This observational study included 314 consecutive patients with chronic residual grade 3+ or 4+ MR at 30 days follow-up post-TAVR, with 104 (33.1%) treated with staged TEER (TEER group) and 210 (66.9%) with medical therapy alone (MT group). The primary composite outcomes were all-cause mortality and heart failure (HF) hospitalization at 2 years. Additional analysis, including changes in MR grade and the New York Association (NYHA) functional classification, and subgroup outcome comparisons based on MR etiology were also conducted.

**Results:** The rate of primary composite outcome was lower in the TEER group than that in the MT group (33.7% vs. 48.1%, p = 0.015). Significant improvement in MR grade and NYHA class was observed in the TEER group after 2 years. Subgroup analysis demonstrated that, among patients with degenerative MR, a lower incidence of composite outcome and HF hospitalization was observed in the TEER group (hazard ratio: 0.35; 95% confidence interval: 0.23–0.53, p < 0.001).

**Conclusions:** Staged TEER post-TAVR was associated with reduced MR and improved clinical outcomes. The clinical significance of MR post-TAVR should be carefully evaluated, and TEER should be considered for patients with significant residual MR, particularly those with degenerative MR.

**What is Known**

- Moderate or severe mitral regurgitation (MR) often accompanies severe aortic stenosis (AS).
- Transcatheter aortic valve replacement (TAVR) is effective for high- or intermediate-risk patients with severe AS.
- Transcatheter edge-to-edge repair (TEER) has shown clinical efficacy for significant MR in high-surgical risk patients.
- Although TAVR can reduce pre-existing MR, significant residual MR is still observed post-TAVR and is associated with adverse cardiac events.

**What the Study Adds:**

- This study systematically evaluates the clinical outcomes of TEER for residual significant MR post-TAVR.
- TEER following TAVR is associated with better clinical outcomes than medical therapy alone, with the TEER group having fewer primary composite events of all-cause mortality and heart failure rehospitalization as well as improved functional status.
- Subgroup analyses based on MR etiology suggest that staged TEER is particularly beneficial in patients with degenerative MR.
- The study highlights the importance of re-evaluating residual MR after TAVR to determine the most effective treatment strategy, emphasizing the potential benefits of staged TEER in improving outcomes for high-risk patients.

## Introduction

Moderate or severe mitral regurgitation (MR) frequently accompanies severe aortic stenosis (AS)^1,2^. Surgical double valve replacement or surgical aortic valve replacement (SAVR) with mitral valve repair has been recommended for severe AS and MR in patients with low or intermediate surgical risk^3,4^. However, simultaneous surgical valve interventions increase morbidity and mortality, particularly in high-risk older patients or those with prior sternotomies^5^.

Transcatheter aortic valve replacement (TAVR) is effective and safe for patients with severe AS at high or intermediate risk for SAVR^6,7^. Indications for TAVR are expanding, even for low-risk patients, with promising outcomes reported in several trials^8,9^. TAVR can reduce pre-existing MR by improving left ventricular function, afterload, diastolic filling, and compliance^10,11^. Nevertheless, significant residual MR is observed post-TAVR, correlating with adverse cardiac events^12,13^.

Transcatheter edge-to-edge repair (TEER) is a minimally invasive option for significant MR in high-surgical risk patients. Multiple studies have demonstrated the clinical efficacy and benefits of TEER with medical therapy for significant MR regardless of etiology^14–17^. TEER holds promise as a treatment for residual significant MR to prevent adverse cardiac events in high-risk TAVR patients. However, its benefits and prognostic impact on residual significant MR post-TAVR remain underevaluated. This study investigated clinical outcomes in patients treated with TEER for residual significant MR observed at 30 days post-TAVR versus outcomes of those who received medical therapy alone.

## Methods

### Study design and patient population

This single-center retrospective observational study conducted at Cedars-Sinai Medical Center enrolled 1,763 consecutive patients aged ≥ 18 years who underwent TAVR for AS between April 2016 and November 2020 (Figure 1). We excluded 24 patients who underwent TEER or transcatheter mitral valve replacement (TMVR) within 30 days. A total of 350 patients were identified with significant residual MR by transthoracic echocardiography (TTE), 30 days post-TAVR follow-up. To control for confounding effects, patients treated with TMVR instead of TEER or those who underwent additional concomitant heart valve replacement during TEER were excluded. Data for nine patients were unavailable due to incomplete records or no follow-up. Finally, our study included 314 patients: 104 (33.1%) underwent TEER with medical therapy post-TAVR (TEER group) and 210 (66.9%) received medical therapy alone (MT group).

**Figure 1.**
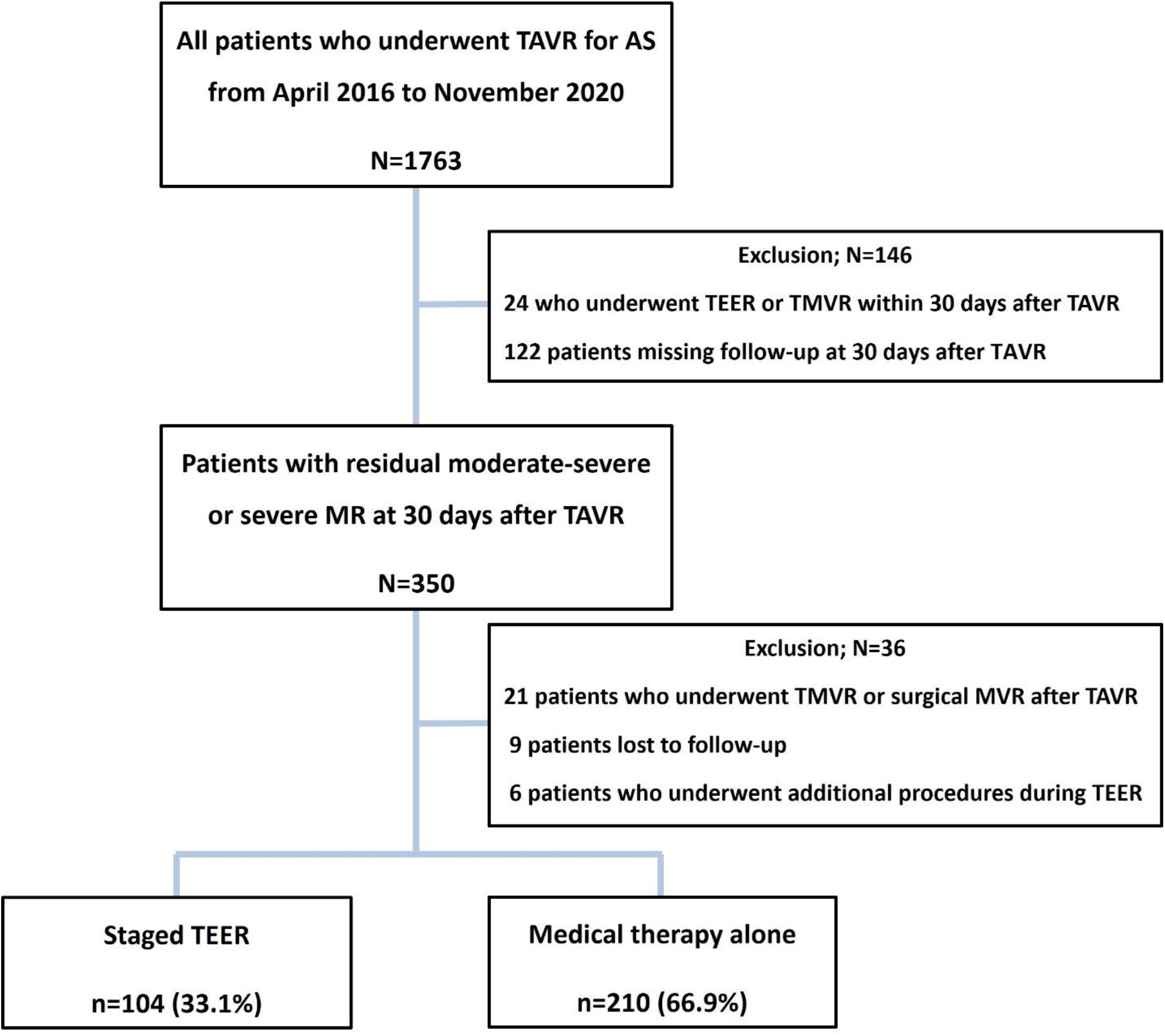
Study flowchart. Number of enrolled patients in the study and their inclusion and exclusion criteria. MR, mitral regurgitation; TAVR, transcatheter aortic valve replacement; TEER, transcatheter mitral edge-to-edge repair; TMVR, transcatheter mitral valve replacement

This study was conducted in accordance with the Declaration of Helsinki and approved by the Institutional Review Board of the Cedars-Sinai Medical Center. All patients provided informed consent.

Patient data were retrospectively obtained from an established interventional cardiology laboratory database at our institution, outpatient visits, and telephone interviews. All patients underwent TTE 30 days post-TAVR as part of a routine post-TAVR echocardiographic assessment, including valvular function evaluation. Follow-up TTE was performed at 6 months, 1 year, and 2 years post-TAVR. MR severity was rated as none-trace (0), mild (1+), moderate (2+), moderate-severe (3+), or severe (4+) based on the American Society of Echocardiography guidelines^18^. Chronic kidney disease (CKD) was defined as an estimated glomerular filtration rate (eGFR) of ≤ 60 mL/min/1.73 m^2^. Anemia was defined as hemoglobin level < 13 mg/dL in men and < 12 mg/dL in women based on World Health Organization criteria^19^.

### Devices and procedures

A multidisciplinary heart team at Cedars-Sinai Medical Center discussed the TEER indication. Procedure details, including device selection and access site, were determined based on preoperative TTE and transesophageal echocardiography. TEER was performed using the MitraClip device (Abbott Vascular, California, USA) according to standard clinical techniques with published guidelines under fluoroscopic and echocardiographic guidance^18^. Procedures were performed under general anesthesia through femoral vein access.

### Outcomes

The primary outcome was a composite outcome of all-cause mortality or heart failure (HF) hospitalization at 2 years after 30 days post-TAVR. Secondary outcomes included all-cause mortality and HF hospitalization. In-hospital complications and clinical assessments, including the New York Heart Association (NYHA) functional class, Kansas City Cardiomyopathy Questionnaire (KCCQ) overall score, and 6-minute walk distance (6 MWD), were also recorded. All clinical outcomes were assessed according to the Mitral Valve Academic Research Consortium criteria^20^. Additional subgroup analyses were performed to explore clinical outcomes based on MR etiology.

### Statistical analyses

Continuous variables are presented as mean ± standard deviation or median and interquartile range (IQR) and categorical variables as numbers and percentages. Between-group differences were evaluated using unpaired Student’s t-test or Mann–Whitney U-test for continuous variables. We used the chi-square or two-tailed Fisher’s exact test for discrete variables and the Wilcoxon signed-rank test for paired ordinal variables (NYHA class and MR grade). Cumulative event rates were estimated using Kaplan–Meier survival analysis, and the log-rank test was used to compare groups. To examine the association between clinical findings and primary outcome, a Cox proportional hazards regression model was developed and expressed as hazard ratios with confidence intervals and p-values, in which, univariate analysis was performed for each potential predictor of 2-year all-cause mortality or HF hospitalization. Only variables with p < 0.05 were incorporated into a stepwise Cox multivariable model. For subgroup analyses, we generated two Cox hazard models: Model 1 adjusted for age and sex. Model 2 included age, sex, and additional clinical variables that showed p < 0.10 (anemia, prior HF hospitalization, and eGFR) at baseline. Two-sided p-values < 0.05 were considered statistically significant. All analyses were performed using SPSS (version 24.0; IBM Corp) and R (version 4.3.2; R Foundation for Statistical Computing).

## Results

### Baseline patient and echocardiographic characteristics

Overall mean age was 79.1 ± 9.5 years (60.2% male) (Table S1). The TEER group exhibited a higher prevalence of anemia, CKD, and NYHA functional class III and IV symptoms than that of the MT group. Baseline echocardiographic characteristics are summarized in Table S2. Overall, 124 patients (39.5%) had degenerative MR (DMR), whereas 190 patients (60.5%) had functional MR (FMR). Table S3 shows procedural demographics of previous TAVR.

### Procedural outcomes and in-hospital TEER complications (Table 1)

The median interval between TEER and TAVR was 48 (IQR, 39–69) days. One clip was used in 47 (45.2%) patients, and the MitraClip G3 system was used most frequently (n = 52, 50.0%). No cases of surgery for failed TEER or tamponade were reported. The in-hospital mortality and reintervention rates before discharge were 1.9% and 2.9%, respectively, and average length of TEER in-hospital stay was 2.04 ± 2.10 days.

**Table 1.** In-hospital outcomes/procedural data for patients with staged TEER performed after 30 days post-TAVR.

|  | TEER<br>(n = 104) |
| --- | --- |
| Procedural data |  |
| Interval from TAVR, days | 48 (39–69) |
| Success implantation | 102 (98.1) |
| Number of clips |  |
| 1 | 47 (45.2) |
| 2 | 42 (40.4) |
| 3 | 15 (14.4) |
| Mitra Clip generations |  |
| MitraClip G2 NT | 31 (29.8) |
| MitraClip G3 NTR/XTR | 52 (50.0) |
| MitraClip G4 NTW/XTW | 21 (20.2) |
| Location of deployed devices |  |
| A1P1 | 6 (5.8) |
| A2P2 | 95 (91.3) |
| A3P3 | 8 (7.7) |
| Procedure time (min) | 116.6 ± 14.6 |
| Fluoroscopy time (min) | 24.9 ± 11.0 |
| Length of hospital stay (days) | 2 (1–4) |
| In-hospital outcomes |  |
| In-hospital death | 1 (1.0) |
| Tamponade | 0 |
| Reintervention | 3 (2.9) |
| Surgery for failed TEER | 0 |
| Single leaflet device attachment | 2 (1.9) |
| Device embolization | 0 |
| Endocarditis | 0 |
| Major bleeding | 3 (2.9) |
| Major vascular complication | 2 (1.9) |
| Stroke | 1 (1.0) |
| Pulmonary thromboembolism | 1 (1.0) |
| Acute kidney injury | 2 (1.9) |
| New onset of atrial fibrillation | 3 (2.9) |

Values presented as mean ± SD, n (%), or median (IQR).
TAVR, transcatheter aortic valve replacement; TEER, transcatheter edge-to-edge repair.

### Clinical outcomes

The median follow-up period was 730 (IQR: 158–763) days. The TEER group exhibited a significantly lower primary composite outcome event rate at 2 years than that of the MT group (Table 2). No significant difference was observed in all-cause mortality at 2 years (11.5% vs. 13.8%; p = 0.57). The observed HF hospitalization rate trended lower at 2 years in the TEER group than that in the MT group (31.7% and 42.9%, respectively, p = 0.057).

**Table 2.** Clinical outcomes at 2 years after 30 days post-TAVR.

|  | Overall<br>(n = 314) | TEER<br>(n = 104) | Medical<br>Therapy<br>(n = 210) | p-value |
| --- | --- | --- | --- | --- |
| Composite outcome | 136 (43.3) | 35 (33.7) | 101 (48.1) | 0.015 |
| All-cause mortality | 41 (13.1) | 12 (11.5) | 29 (13.8) | 0.57 |
| HF hospitalization | 123 (39.2) | 33 (31.7) | 90 (42.9) | 0.057 |

Values presented as n (%).
HF, heart failure; TAVR, transcatheter aortic valve replacement; TEER, transcatheter edge-to-edge repair

### Clinical functional measurements

The NYHA functional class improved in both groups from baseline to 6 months, with the TEER group showing marked change (p < 0.001) (Figure S1). Although the change in NYHA class at 2 years remained significantly different from baseline in the TEER group, no such difference was noted for the MT group. Similarly, KCCQ score and 6 MWD in the TEER group improved from baseline to 1 and 2 years (Figure S2A, S2B).

### Echocardiographic follow-up data

In the TEER group, MR grade at 6 months significantly improved relative to baseline (p < 0.001; Figure 2), and was sustained at 1 and 2 years. An MR reduction trend in the MT group was observed after 6 months, without statistical significance (p = 0.052). The change in left ventricular ejection fraction (LVEF) from baseline (before TEER) to 6-month follow-up was comparable between groups (p = 0.16, Table S4).

**Figure 2.**
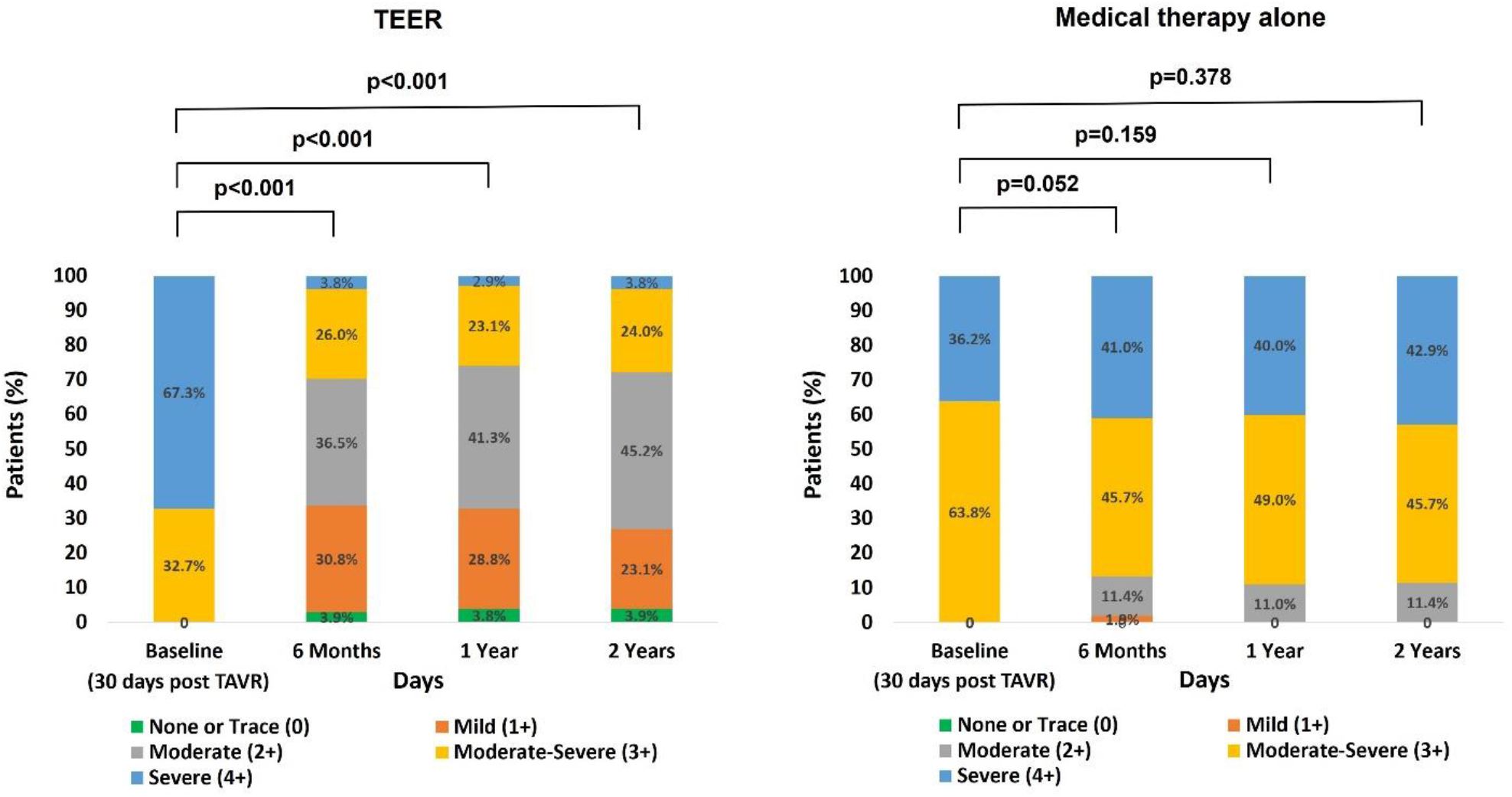
Residual MR post-procedure during follow-up period. Changes in mitral regurgitation (MR) severity from baseline to follow-up in the TEER group (left) and medical therapy group (right). Wilcoxon signed-rank test was used to determine the statistical significance of these changes. MR, mitral regurgitation; TAVR, transcatheter aortic valve replacement; TEER, transcatheter mitral edge-to-edge repair

### Predictors and primary outcome subgroup analysis

Multivariable analysis revealed lower risk of all-cause mortality or HF hospitalization in the TEER group than that in the MT group (HR: 0.35; CI: 0.23–0.53, p < 0.001) (Table 3). The MR multivariable Cox proportional hazard model revealed age, NYHA functional class IV symptoms, CKD, LVEF, and residual severe as independent predictors of the composite outcome. Incidences of the primary composite outcome in various major subgroups are shown in Figure 3. Furthermore, patients were divided into groups based on MR etiology (DMR and FMR), and clinical outcomes were compared between the TEER and MT groups based on MR etiology. Baseline characteristics are shown in Table S5. In patients with DMR, a lower incidence of the composite outcome and HF hospitalization was observed in the TEER group than that in the MT group (Figure 4A, 4C), with no significant inter-group difference in all-cause mortality (Figure 4B). In patients with FMR, no significant differences were noted between the treatment groups regarding composite outcome, all-cause mortality, and HF hospitalization (Figure 4D–F). However, patients with FMR in the TEER group showed significantly lower HR for the composite outcome than that of those in the MT group after adjusting for age, sex, anemia, prior HF hospitalizations, and eGFR (Model 2: HR 0.59, 95% CI, 0.37–0.99; p = 0.047) (Table S6).

**Figure 3.**
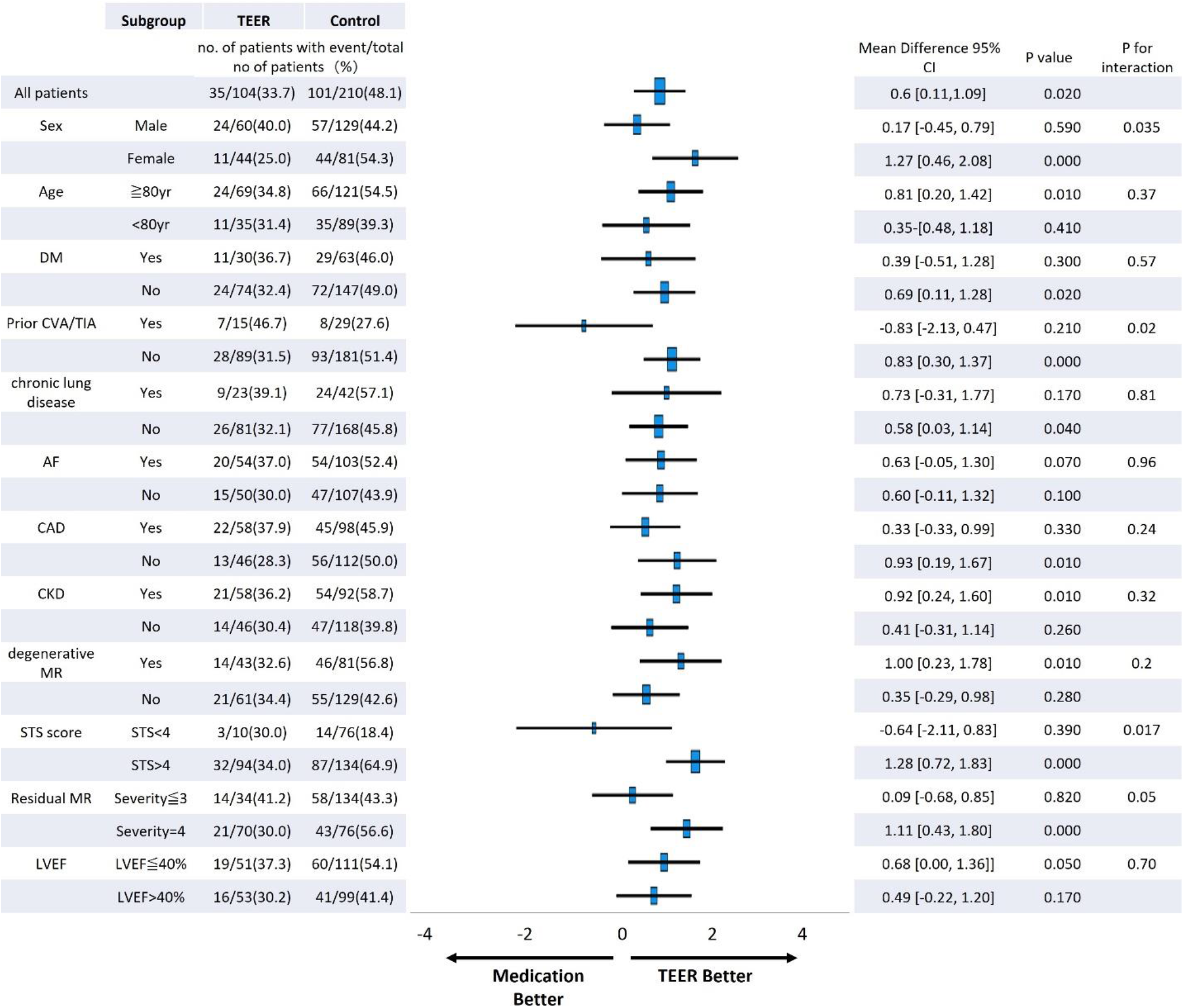
Subgroup analyses for primary composite outcome of all-cause mortality or heart failure hospitalization. Blue boxes represent mean difference, with horizontal lines illustrating 95% confidence intervals. Dimensions of each box are proportionally aligned with the size of its relevant subgroup. TEER, transcatheter edge-to-edge repair; DM, diabetes mellitus; CVA, cerebrovascular accident; TIA, transient ischemic attack; AF, atrial fibrillation; CAD, coronary artery disease; CKD, chronic kidney disease; MR, mitral regurgitation; NYHA; STS, Society of Thoracic Surgeons; LVEF, left ventricular ejection fraction.

**Figure 4.**
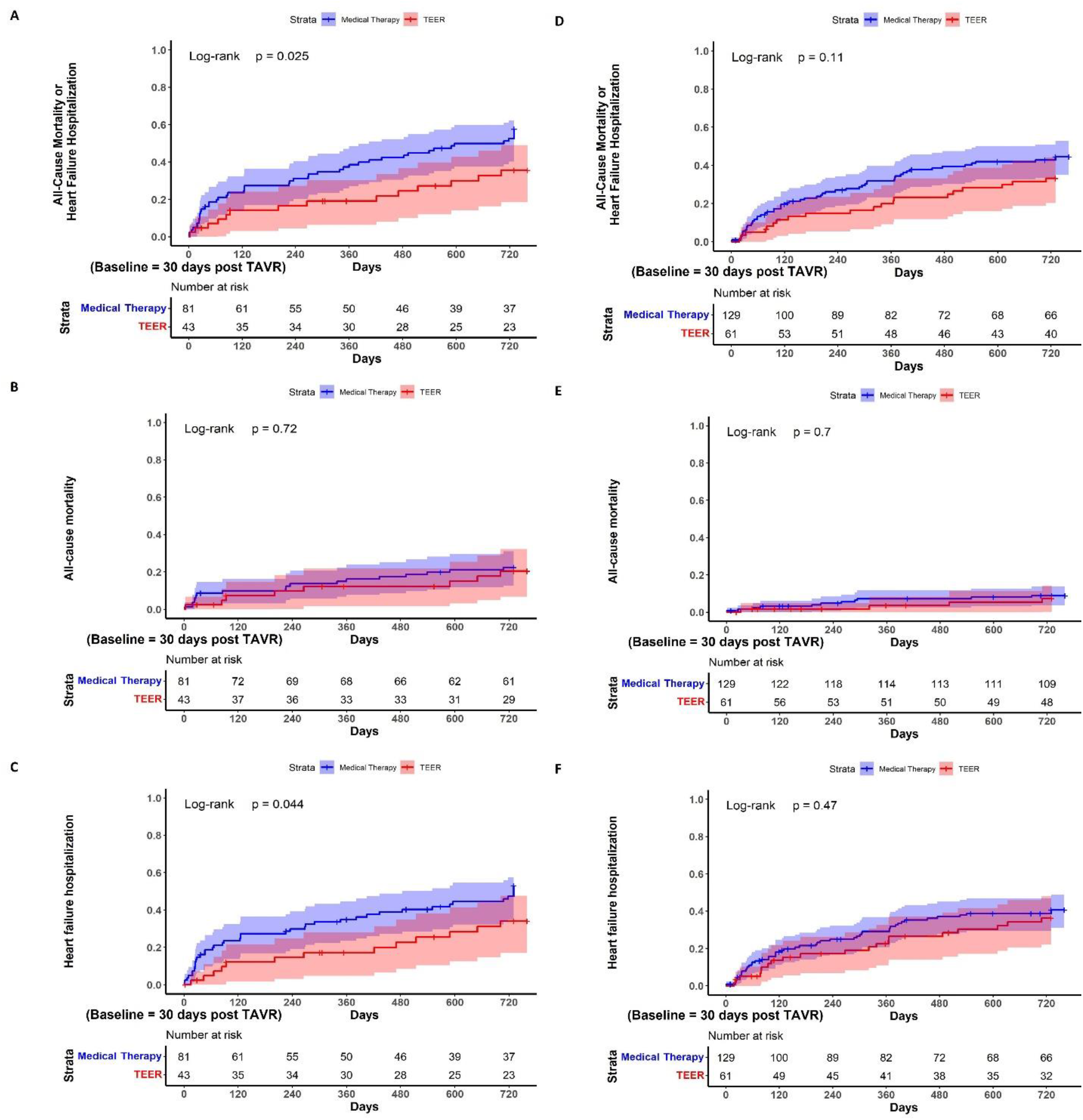
Kaplan–Meier curves for patients with degenerative mitral regurgitation. (A) Primary composite outcome (all-cause mortality or heart failure hospitalization), (B) all-cause mortality, and (C) heart failure hospitalization through 2 years in patients with degenerative mitral regurgitation. Kaplan–Meier curves shown for (D) the primary composite outcome (all-cause mortality or heart failure hospitalization), (E) all-cause mortality, and (F) heart failure hospitalization through 2 years in patients with functional mitral regurgitation. Event rates were calculated using Kaplan–Meier analysis and compared using the log-rank test. TAVR, transcatheter aortic valve replacement, TEER, transcatheter mitral edge-to-edge repair

**Table 3.** Predictors of all-cause mortality or heart failure hospitalization.

|  | Univariable model |  | Multivariable model |  |
| --- | --- | --- | --- | --- |
|  | HR (95% CI) | p value | HR (95% CI) | p value |
| TEER | 0.65 (0.44–0.96) | 0.027 | 0.35 (0.23–0.53) | < 0.001 |
| Age, years | 1.03 (1.01–1.05) | 0.006 | 1.04 (1.02–1.06) | 0.001 |
| Male | 1.06 (0.75–1.49) | 0.75 | 0.85 (0.61–1.22) | 0.86 |
| BMI | 1.01 (0.98–1.04) | 0.51 |  |  |
| NYHA functional Class = IV | 2.56 (1.82–3.60) | < 0.001 | 2.93 (2.05–4.18) | < 0.001 |
| STS score, per 1 point | 1.04 (1.02–1.06) | < 0.001 |  |  |
| Chronic kidney disease | 1.45 (1.04–2.04) | 0.03 | 1.61 (1.14–2.28) | 0.007 |
| Peripheral vascular disease | 0.59 (0.31–1.13) | 0.11 |  |  |
| Prior stroke | 0.66 (0.39–1.13) | 0.13 |  |  |
| Chronic lung disease | 1.34 (0.91–1.98) | 0.14 |  |  |
| Coronary artery disease | 1.00 (0.72–1.40) | 0.99 |  |  |
| Prior atrial fibrillation | 1.27 (0.91–1.78) | 0.17 |  |  |
| Prior pacemaker/ICD | 1.5 (0.79–1.66) | 0.47 |  |  |
| Anemia | 1.21 (0.86–1.70) | 0.28 |  |  |
| Beta-blocker | 1.3 (0.87–1.93) | 0.19 |  |  |
| ACE inhibitor/ARB | 0.89 (0.64–1.25) | 0.51 |  |  |
| Residual severe MR | 1.48 (1.06–2.08) | 0.022 | 1.70 (1.12–2.42) | 0.003 |
| Tricuspid regurgitation ≥ moderate | 1.30 (0.93–1.82) | 0.13 | | |
| LVEF, per increase 10% | 0.88 (0.79–0.98) | 0.017 | 0.86 (0.77–0.96) | 0.009 |
| LV end-diastolic diameter (cm) | 1.08 (0.92–1.26) | 0.35 |  |  |
| Pulmonary systolic artery pressure (mmHg) | 1.01 (0.99–1.03) | 0.32 |  |  |

ACE-I, angiotensin-converting enzyme inhibitors; ARBs, angiotensin-receptor blockers; BMI,
body mass index; CI, confidence interval; HR, hazard ratio; ICD, implantable cardioverter-
defibrillator; LVEF, left ventricular ejection fraction; MR, mitral regurgitation; NYHA, New
York Heart Association; STS, Society of Thoracic Surgeons; TEER, transcatheter edge-to-edge repair.

## Discussion

Our study is the first to compare clinical and echocardiographic TEER outcomes with those of medical therapy alone in patients with significant residual MR post-TAVR and complete additional subgroup analyses based on MR etiology. We found low in-hospital complication rates in the TEER group, reaffirming the safety of TEER post-TAVR. TEER post-TAVR was associated with better clinical outcomes compared to medical therapy alone, with the TEER group having fewer primary composite events and improved functional status. Finally, the clinical benefit of staged TEER post-TAVR was more substantial in patients with residual DMR than in those with FMR.

Additional mitral valve intervention for significant residual MR post-TAVR can reduce mortality^21^. However, previous studies included patients who underwent TMVR, had residual MR grade ≤ 2+, were treated more than a decade ago, or received early staged TEER post-TAVR. Conversely, our study enrolled patients with residual MR grade ≥ 3+ undergoing staged TEER using a new-generation mitral clip system. Before considering staged TEER, our patients were re-evaluated for MR severity by TTE (post-TAVR). Additionally, patients who underwent TEER within 30 days of TAVR were excluded to prevent including patients treated prematurely for MR that could potentially resolve unassisted post-TAVR^22^. Patzelt et al. described that mitral valve anatomy in MR patients is altered due to TAVR^23^. Lastly, a higher complication rate and longer hospital stays were reported with concomitant TAVR and TEER compared with staged TEER post-TAVR^24^, underscoring the importance of re-evaluating residual MR post-TAVR to determine optimal strategies for mitral valve intervention, avoiding routine concomitant or early-staged TEER post-TAVR.

The in-hospital complication rate for staged TEER was relatively low in our study, consistent with previous randomized study findings^16,25^. Furthermore, compared to medical therapy alone, staged TEER was associated with a lower incidence of the primary composite outcome and greater MR reduction. Although the between-group difference in HF hospitalization incidence did not reach statistical significance (p = 0.066), the trend suggested TEER may reduce HF hospitalization risk. Several factors may explain the favorable outcome of staged TEER post-TAVR. Recently, notable progress has been observed in the development of innovative TEER devices. We used MitraClip G2 NT and new-generation mitral clips, including G3 NTR/XTR and G4 NTW/XTW for TEER; the first-generation mitral clip was not used, and 70% of patients received new-generation clips. The MitraClip EXPAND Study demonstrated that mitral valve repair with MitraClip NTR/XTR is associated with a lower rate of leaflet adverse events than that of those previously reported using older-generation devices^26^. Additionally, the MitraClip G4 system for TEER was associated with greater MR reduction^27^. Therefore, the use of new-generation devices may contribute to improved patient outcomes. Furthermore, increased institutional experience in the TEER procedure may correlate with improved procedural outcomes and shorter hospital stays^28^. In the present study, the hospital stay period for patients who underwent staged TEER after prior TAVR was notably short, indicating the benefit and feasibility of staged TEER post-TAVR for avoiding a functional decline in older or frail patients.

Staged TEER drastically improved the NYHA class, KCCQ score, and 6 WMD, indicating that TEER was an effective treatment option for symptomatic patients with residual MR post-TAVR. The NYHA class and 6 WMD improved at 1 year in patients receiving medical therapy alone, but was not sustained at 2 years; our findings emphasize staged TEER as a beneficial intervention for symptom improvement in these patients.

We demonstrated the efficacy of staged TEER after 30 days post-TAVR according to individual MR etiologies, with subgroup analysis showing interesting results. Significant differences in composite outcomes and HF hospitalization rates were observed between the TEER and MT groups in patients with DMR. However, no significant differences were noted in those with FMR. Furthermore, TAVR is unlikely to improve the magnitude of MR related to degenerative anatomical pathology, and TEER may be an effective treatment by directly reducing MR caused by DMR. Conversely, TAVR can reduce FMR with improvements in afterload, LVEF, diastolic function, or reverse remodeling^10,29^. Although the decrease in afterload and LV pressure post-TAVR may result in early MR improvement, the effect of reverse remodeling leading to FMR improvement could take longer. Toggweiler et al. documented improvements in LVEF and left ventricular end-diastolic diameter among patients with moderate or severe MR undergoing TAVR at the 1-year follow-up. Patients with FMR were more prone to MR reductions at the 1-year follow-up assessment than those with DMR^30^. Although our study focused on patients with significant residual MR at 30 days post-TAVR, FMR may have improved long-term, even in patients receiving medical therapy alone post-TAVR, potentially leading to favorable outcomes without the need for TEER. However, after adjustment for age, sex, anemia, prior HF hospitalization, and eGFR, staged TEER for FMR was associated with lower rates of the composite outcomes. This finding highlights potential benefits of staged TEER in select patients with FMR who have achieved optimal medical management. Considering the long-term improvement in FMR post-TAVR, a prolonged waiting period before performing staged TEER post-TAVR could result in more positive outcomes. Further studies are necessary to determine the optimal timing and criteria for patient selection when TEER is considered for patients with significant FMR post-TAVR.

Staged TEER reduced the rate of HF hospitalization and all-cause mortality, with reduction in MR grades. The results clearly show that TEER is a viable option for treating residual MR in high-risk symptomatic patients, even post-TAVR. Since TAVR has continued to evolve to meet the expanding indications and growing use in routine clinical practice^8^, the number of patients with significant MR post-TAVR may increase with time. The present study provides novel insights into the clinical outcomes of staged TEER post-TAVR for patients with significant residual MR, offering perspectives for future management of patients with both AS and MR.

This study has limitations. First, it was a single-center study with a relatively small sample size. Second, the study’s retrospective and observational design may have led to selection bias for TEER. Patients who underwent staged TEER post-TAVR tended to have higher risks than those who received medical therapy alone. Although multivariable analysis was conducted to adjust for confounding factors, we could not exclude potential factors affecting clinical outcomes. Moreover, TEER may not be feasible for some patients receiving medical therapy alone due to anatomical issues. Accordingly, randomized, double-blind studies are needed to avoid potential bias. Third, the timing of TEER varied among patients, possibly impacting their outcomes. Further studies should determine the optimal timing of intervention for improved outcomes. Lastly, we assessed outcomes for up to 2 years; however, a longer follow-up period is required to clarify accurate outcomes for each treatment.

In conclusion, staged TEER post-TAVR is safe and effective for significant residual MR in patients who underwent TAVR. Staged TEER may improve clinical outcomes at 2 years with improved symptoms and lower in-hospital complication rates than those of medical therapy alone, especially in patients with DMR. Randomized controlled trials with larger sample sizes and longer follow-up periods are needed to confirm further our results of staged TEER for residual significant MR post-TAVR.

## Data Availability

The data that support the findings of this study are available upon reasonable request from the corresponding author.

## Acknowledgments

None

## Non-standard Abbreviations and Acronyms

AS: Aortic stenosis
CKD: Chronic kidney disease
DMR: Degenerative mitral regurgitation
FMR: Functional mitral regurgitation
HF: Heart failure
IQR: Interquartile range
KCCQ: Kansas City Cardiomyopathy Questionnaire
LVEF: Left ventricular ejection fraction
MR: Mitral regurgitation
NYHA: New York Heart Association
STS: Society of Thoracic Surgeons
SAVR: Surgical aortic valve replacement
TAVR: Transcatheter aortic valve replacement
THV: Transcatheter heart valve
TEER: Transcatheter edge-to-edge repair
TMVR: Transcatheter mitral valve replacement
TTE: Transthoracic echocardiogram
6 MWD: 6-minute walk distance

## Supplemental Material

Tables S1–S6

Figures S1, S2

